# Incidence Measures for Schizophrenia among Commercial Health Insurance and Medicaid Enrollees

**DOI:** 10.1101/2023.02.24.23286405

**Authors:** Molly T. Finnerty, Atif Khan, Kai You, Rui Wang, Gyojeong Gu, Deborah Layman, Qingxian Chen, Noémie Elhadad, Shalmali Joshi, Paul S. Appelbaum, Todd Lencz, Sander Markx, Steven A. Kushner, Andrey Rzhetsky

**Affiliations:** Office of Population Health and Evaluation, NYC Field Office, New York State Office of Mental Health, New York, NY; Department of Child and Adolescent Psychiatry, New York University Grossman School of Medicine, New York, NY; Department of Medicine, and Institute of Genomics and Systems Biology, University of Chicago, Chicago, IL; Department of Biomedical Informatics, Columbia University New York, NY; Department of Psychiatry, Columbia University College of Physicians & Surgeons; New York State Psychiatric Institute, New York, NY; Institute of Behavioral Science, Feinstein Institutes for Medical Research, Manhasset, NY; Department of Psychiatry, Zucker School of Medicine at Hofstra/Northwell, Hempstead, NY; Institute for Genomics and Systems Biology, University of Chicago, Chicago, IL

**Keywords:** schizophrenia, new onset, incidence, prevalence, Medicaid, MarketScan, lifespan

## Abstract

**Objective:** To examine and compare the prevalence, incidence, and specificity and positive predictive value of incidence measures in a national commercial insurance database and a state Medicaid database, for different age groups over the life span.

**Methods:** This cross-sectional study examined the annual prevalence of schizophrenia in 2019 in two databases: a) MarketScan, a United States national commercial insurance database (*N* = 16,365,997), and b) New York State Medicaid program (*N* = 4,414,153). A subset of these individuals with 10 years of continuous insurance prior to 2019 (2009-2018) was used to assess incidence and to create a reference standard for incidence measure testing, including positive predictive value and specificity, by age group. Continuous enrollment was required for all years of study (45-day gap allowance per year, and one or more health care services annually to ensure opportunity to be diagnosed).

**Results:** The prevalence of schizophrenia in 2019 was 0.13% in MarketScan, and 2.13% in Medicaid. Prevalence was highest in MarketScan among individuals aged 21-25 years. In contrast, prevalence in Medicaid increased over the lifespan, with the peak among those 56-60 years of age. The incidence of new-onset schizophrenia in 2019 was 0.07% in MarketScan and 0.19% in Medicaid and peaked in young adulthood (26-30 years of age) in both databases (0.16% and 0.40%, respectively). Positive predictive value (PPV) for the incidence measures was higher for women than men and increased with longer lookback periods prior to the first schizophrenia diagnosis in 2019. Using a one-year lookback period, the PPV for those ≤35 years of age was 79% in MarketScan and 51% in Medicaid; a ≥95% PPV was achieved with a two-year lookback in MarketScan, and a seven-year lookback period in Medicaid. PPVs were also higher in younger age groups; in Medicaid a two-year lookback period yielded a PPV of 68% for those ≤35 years, but was 94% for those ≤20 years. Specificity was high even for the total population with one year of observation prior to the first schizophrenia diagnosis in 2019, and was higher in Medicaid (92%) than in MarketScan (80%), increasing further with longer lookback periods in both databases.

**Conclusions:** The prevalence of schizophrenia is over 10-fold higher, and the incidence two-fold higher, in the New York State Medicaid population compared to the MarketScan national commercial insurance database. Within each database, prevalence and incidence varied by sex, age, and duration of insurance. Yet despite these differences, accurate measures of incidence of new-onset schizophrenia could be constructed with high specificity and positive predictive values.

## INTRODUCTION

Schizophrenia is a severe mental health disorder marked by cognitive and emotional disturbances that significantly impact an individual’s social functioning (American Psychiatric Association, 2013). Estimates of lifetime prevalence of schizophrenia have varied widely due to the heterogeneity of the disorder, as well as the study methods used, including the design, prevalence period type, data source and data quality (Simeone, Ward, Rotella, Collines, & Windisch, 2015). Estimates of the prevalence of schizophrenia in the United States vary from 0.3 - 1.6% in community-based samples, 1.02% in uninsured samples, 0.13% in commercial insurance samples, and 1.66% - 5.11% in Medicaid (Kessler et al., 2005; Pilon et al., 2021; Wu et al., 2006). Prevalence estimates have often relied upon community-based data estimates from over two decades ago (Kessler et al., 2005), while more recent studies have shown considerable promise leveraging pooled datasets (Pilon et al., 2021).

Surveillance methods in the United States face several challenges. The United States does not have a national health registry database, and national studies of community samples are costly. Moreover, occupational loss, institutionalization, and downward socioeconomic trajectories have greatly complicated naturalistic prospective longitudinal observational studies of individuals with schizophrenia. Researchers and governments have relied on large administrative claims databases, such as Medicaid, to conduct surveillance and epidemiological studies. Medicaid includes a large proportion of individuals with schizophrenia; however, it may underestimate incidence and prevalence when used alone (Pilon et al., 2021). Accordingly, using multiple independent databases may expand and diversify the population observed, allow for comparison of the prevalence and incidence by insurance type, and support testing and refinement of incidence measures to increase generalizability of assessment methods.

Given the chronic nature of schizophrenia, it is also important to examine prevalence and incidence across the lifespan for age-related differences to avoid underestimating prevalence and incidence. Accuracy and ongoing monitoring of prevalence and incidence are important to understand the scope of the burden of schizophrenia across the lifespan and to optimize interventions at specific developmental ages and within defined subgroups most in need of resources.

The current study compares the prevalence, incidence, and specificity and predictive value of incidence measures with varying lookback periods using two administrative databases in the United States. We hypothesized that a national commercial insurance database of employed individuals and their families (MarketScan) would exhibit a lower prevalence and higher proportion of incident to prevalent cases, due to an expected disproportionate loss of employer provided commercial insurance for the schizophrenia population. We also hypothesized that the Medicaid population would have higher prevalence rates as individuals with schizophrenia have a higher burden of socioeconomic risk factors, in addition to the tendency to transition from commercial insurance to Medicaid after developing schizophrenia. We examined the specificity and positive predictive value of incidence measures in these two databases across the lifespan, with the goal of addressing a critical gap to inform future schizophrenia research and policy development using large disparate databases.

## METHODS

### Data Sources

This cross-sectional study used two databases: 1) MarketScan, a proprietary deidentified national research database that comprises largely individuals covered under employer-sponsored commercial insurance, including employed individuals, their spouses, and dependents; and 2) New York State Medicaid, a large state public insurance program.

### Study Populations

Total population included all individuals in the MarketScan and Medicaid databases in 2019 with 12 months of continuous insurance (allowing for a 45-day gap), and one or more health services (inpatient or outpatient) during 2019. The 10-year continuously insured cohort was the subset of the total population that had continuous insurance in the 10 years prior to 2019 (1/1/2009-12/31/2018), with 12 months of coverage in each year, and no more than a 45-day gap in any year. To ensure that individuals had an opportunity to be assessed, one or more health service visits per year was an additional criterion for study inclusion.

### Definitions and Measures

Demographic characteristics included age (as of 12/31/2019), constructed age cohort groups (0-10, 11-15, 16-20, 21-25, 26-30, 31-35, 36-40, 41-45, 46-50, 51-55, 56-60, 61-64, 65+ years of age), a ≤35 years of age group, and sex (male/female). Race and ethnicity (available only in Medicaid database) were categorized as Hispanic or Latinx (any race), or non-Hispanic White, Black, Asian, American Indian, Multiracial, or Unknown. Prevalent cases of schizophrenia were defined as those with two or more diagnoses of schizophrenia or schizoaffective disorder (ICD-9: 295.x; ICD-10: F20, F25; excluding schizophreniform: 295.4x, F20.81) in 2019. Incident (new-onset) cases of schizophrenia were defined as the subset of individuals in the 10-year continuously insured cohort who met criteria for schizophrenia in 2019 and had no prior schizophrenia diagnoses in the 10 years prior (1/1/2009-12/31/2018).

### ANALYSIS

The prevalence of schizophrenia in 2019 was calculated for the total population, and for the 10-year continuously insured cohorts in MarketScan and in Medicaid. The incidence of schizophrenia (new-onset in 2019) was calculated for the 10-year continuously insured cohorts only. To identify new-onset cases of schizophrenia, ideally an individual’s lifelong medical record would be reviewed, but no such database exists in the United States. We used a 10-year lookback period without any prior evidence of schizophrenia as the reference standard in Medicaid and MarketScan for defining incident cases of schizophrenia. Any case of schizophrenia diagnosed in 2019 that had no evidence of a schizophrenia diagnosis in the decade prior (1/1/2009-12/31/2018), was considered an incident case.

We examined the accuracy of using shorter lookback periods to identify incident cases of schizophrenia, since using a 10-year lookback period greatly reduces the size of the study population to those with 10 years of continuous observation in the database, and a 10-year lookback may not be feasible for some studies or databases. A variety of methods have been reported in the literature to assess incidence, ranging from lifetime review to a period as short as one-year prior to the index schizophrenia date (Pilon et al., 2021), but little data is available on the accuracy of these estimates to inform methodological decisions. Schizophrenia is a chronic condition, and it may be expected that for those engaged in care a single year or two of lookback may capture nearly all of the cases that would be identified in a much longer or lifelong review. However, if there is diagnostic uncertainty in the early years of illness, or if some individuals are diagnosed but have long gaps in engagement in treatment, then shorter lookback periods will miss prior diagnoses of schizophrenia. We examined the accuracy of measures of new-onset of schizophrenia, testing lookback periods of “x” duration (1, 2, 3, 4, 5, 6, 7, 8 and 9 years), compared to a reference standard of a 10-year lookback period prior to the first observed diagnosis of schizophrenia in 2019 (Figure 1). We calculated the positive predictive value (PPV) and specificity (Trevethan, 2017) of incidence measures of new-onset schizophrenia using sequentially longer lookback periods. Sensitivity and negative predictive value testing were not required since both were 100% for all values of “x” years of lookback due to the absence of false negatives; all prior diagnosis of schizophrenia identified within “x” years, will also be identified with a longer lookback period.

**Figure 1.**
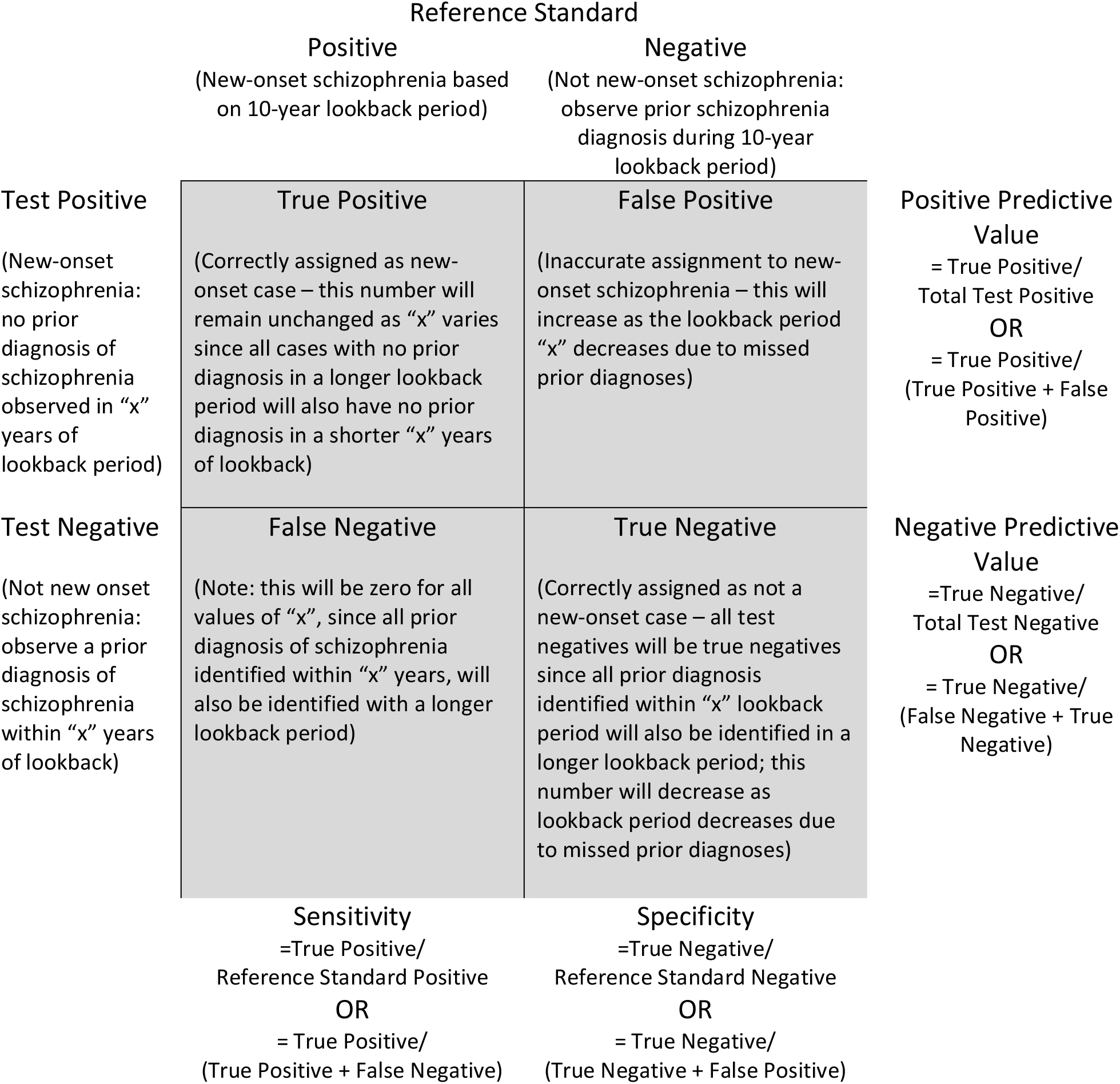
Definitions of positive predictive value and specificity - adapting accuracy testing to incidence measures of new-onset schizophrenia.

The New York State Office of Mental Health IRB approved the study protocol. In addition, the University of Chicago IRB approved their use of MarketScan data as IRB-exempt.

## RESULTS

### Population Characteristics

The characteristics of the 2019 total population and 10-year continuously insured cohorts for MarketScan (*N* = 16,365,997 and *n* = 951,173, respectively) and Medicaid (*N* = 4,414,153 and *n* = 785,088, respectively) are summarized in Table 1. The mean age (± SD) was older in MarketScan (38.1 ± 20.2 years) than in Medicaid (33.5 ± 24.3 years). The proportion of females was similar in both databases (MarketScan 54.1%, Medicaid 55.7%), and increased in the 10-year insured cohort (MarketScan 62%, Medicaid 61.1%). In Medicaid, the Multiracial group was the largest (31.5%), followed by White (26.5%), Black (15.8%), Hispanic or Latinx (12.2%), Asian (9.6%), and American Indian (0.3%) race and ethnicity groups.

**Table 1.**
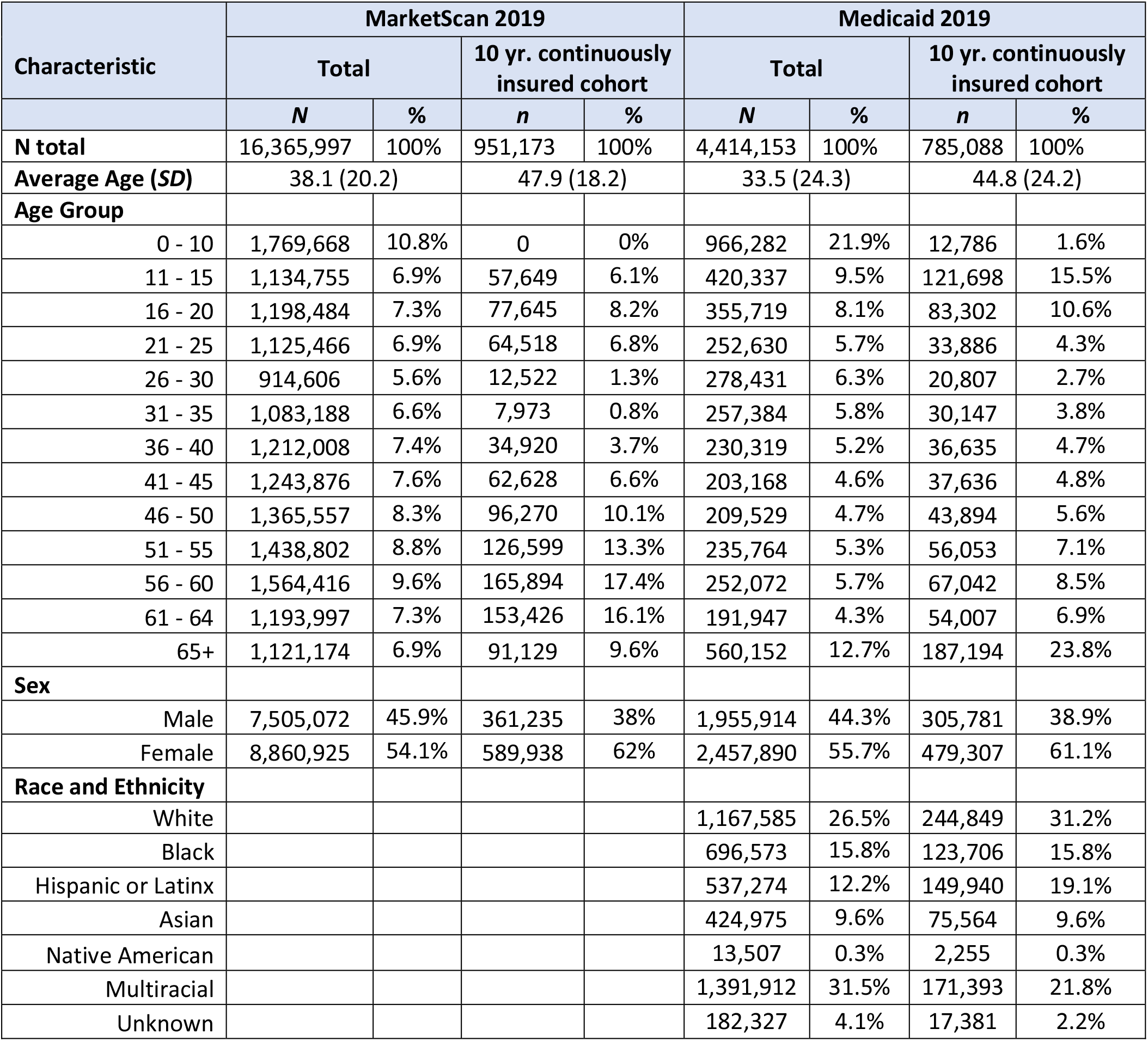
Characteristics of insured individuals in MarketScan and Medicaid in 2019 for total population and 10-years continuously insured cohorts.

| Characteristic | MarketScan 2019 |  |  |  | Medicaid 2019 |  |  |  |
| --- | --- | --- | --- | --- | --- | --- | --- | --- |
|  | Total |  | 10 yr. continuously insured cohort |  | Total |  | 10 yr. continuously insured cohort |  |
|  | *N* | % | *n* | % | *N* | % | *n* | % |
| **N total** | 16,365,997 | 100% | 951,173 | 100% | 4,414,153 | 100% | 785,088 | 100% |
| **Average Age (*SD*)** | 38.1 (20.2) |  | 47.9 (18.2) |  | 33.5 (24.3) |  | 44.8 (24.2) |  |
| **Age Group** |  |  |  |  |  |  |  |  |
| 0 - 10 | 1,769,668 | 10.8% | 0 | 0% | 966,282 | 21.9% | 12,786 | 1.6% |
| 11 - 15 | 1,134,755 | 6.9% | 57,649 | 6.1% | 420,337 | 9.5% | 121,698 | 15.5% |
| 16 - 20 | 1,198,484 | 7.3% | 77,645 | 8.2% | 355,719 | 8.1% | 83,302 | 10.6% |
| 21 - 25 | 1,125,466 | 6.9% | 64,518 | 6.8% | 252,630 | 5.7% | 33,886 | 4.3% |
| 26 - 30 | 914,606 | 5.6% | 12,522 | 1.3% | 278,431 | 6.3% | 20,807 | 2.7% |
| 31 - 35 | 1,083,188 | 6.6% | 7,973 | 0.8% | 257,384 | 5.8% | 30,147 | 3.8% |
| 36 - 40 | 1,212,008 | 7.4% | 34,920 | 3.7% | 230,319 | 5.2% | 36,635 | 4.7% |
| 41 - 45 | 1,243,876 | 7.6% | 62,628 | 6.6% | 203,168 | 4.6% | 37,636 | 4.8% |
| 46 - 50 | 1,365,557 | 8.3% | 96,270 | 10.1% | 209,529 | 4.7% | 43,894 | 5.6% |
| 51 - 55 | 1,438,802 | 8.8% | 126,599 | 13.3% | 235,764 | 5.3% | 56,053 | 7.1% |
| 56 - 60 | 1,564,416 | 9.6% | 165,894 | 17.4% | 252,072 | 5.7% | 67,042 | 8.5% |
| 61 - 64 | 1,193,997 | 7.3% | 153,426 | 16.1% | 191,947 | 4.3% | 54,007 | 6.9% |
| 65+ | 1,121,174 | 6.9% | 91,129 | 9.6% | 560,152 | 12.7% | 187,194 | 23.8% |
| **Sex** |  |  |  |  |  |  |  |  |
| Male | 7,505,072 | 45.9% | 361,235 | 38% | 1,955,914 | 44.3% | 305,781 | 38.9% |
| Female | 8,860,925 | 54.1% | 589,938 | 62% | 2,457,890 | 55.7% | 479,307 | 61.1% |
| **Race and Ethnicity** |  |  |  |  |  |  |  |  |
| White |  |  |  |  | 1,167,585 | 26.5% | 244,849 | 31.2% |
| Black |  |  |  |  | 696,573 | 15.8% | 123,706 | 15.8% |
| Hispanic or Latinx |  |  |  |  | 537,274 | 12.2% | 149,940 | 19.1% |
| Asian |  |  |  |  | 424,975 | 9.6% | 75,564 | 9.6% |
| Native American |  |  |  |  | 13,507 | 0.3% | 2,255 | 0.3% |
| Multiracial |  |  |  |  | 1,391,912 | 31.5% | 171,393 | 21.8% |
| Unknown |  |  |  |  | 182,327 | 4.1% | 17,381 | 2.2% |

### Prevalence

The prevalence of schizophrenia in 2019 for the total MarketScan and Medicaid populations and the 10-year continuously insured cohorts is summarized in Table 2. The prevalence of schizophrenia in the total population was higher in Medicaid (2.13%, *n* = 94,153 of 4,414,153) than in MarketScan (0.13%, *n* = 21,963 of 16,365,997). In both databases the prevalence was higher among the subsample of individuals with 10-years of continuous insurance prior to 2019, but this increase was more marked in Medicaid (5.74% vs. 2.13%) than in MarketScan (0.16% vs. 0.13%). Males had a higher prevalence than females (Medicaid: 2.8% vs. 1.6%; MarketScan: 0.14% vs. 0.13%) in the total population, as well as in the 10-year continuously insured cohort (Medicaid: 7.89% vs. 4.37%; MarketScan: 0.18% vs. 0.14%).

**Table 2.** Prevalence of schizophrenia in MarketScan and Medicaid (2019)

| **Population** |  | **Total** | **Male** | **Female** | **≤35 years** |
| --- | --- | --- | --- | --- | --- |
| Medicaid Total | Total | 4,414,153 | 1,955,914 | 2,457,890 | 1,653,261 |
|  | Schizophrenia | 94,153 | 54,829 | 39,324 | 20,627 |
|  | **Prevalence** | **2.13%** | **2.80%** | **1.60%** | **1.25%** |
| Medicaid 10-year<br>Continuously<br>Insured | Total | 785,088 | 305,781 | 479,307 | 302,619 |
|  | Schizophrenia | 45,068 | 24,130 | 20,938 | 4,687 |
|  | **Prevalence** | **5.74%** | **7.89%** | **4.37%** | **1.55%** |
| MarketScan Total | Total | 16,365,997 | 7,505,072 | 8,860,925 | 5,662,318 |
|  | Schizophrenia | 21,963 | 10,169 | 11,794 | 8,183 |
|  | **Prevalence** | **0.13%** | **0.14%** | **0.13%** | **0.14%** |
| MarketScan 10-year<br>Continuously<br>Insured | Total | 95,1173 | 361,235 | 589,938 | 220,307 |
|  | Schizophrenia | 1,519 | 664 | 855 | 457 |
|  | **Prevalence** | **0.16%** | **0.18%** | **0.14%** | **0.21%** |

The prevalence of schizophrenia by age group for the total population and 10-year continuously insured cohort is presented in Figure 2. The prevalence of schizophrenia in the Medicaid population increased over the course of the lifespan up to age 56-60 years (total population: 5.0%; 10-year continuously insured cohort; 11.3%), consistent with a chronic condition in which individuals preferentially acquire and continuously maintain Medicaid insurance. MarketScan prevalence peaks at a younger age in the total population (21-25 years-old, 0.25%) and among those with 10 years of continuous insurance (26-30 years-old, 0.78%), consistent with a tendency for individuals who develop schizophrenia to lose commercial insurance when they can no longer be covered on their parents’ policy (United States Affordable Care Act, maximum age 26 years-old).

**Figure 2.**
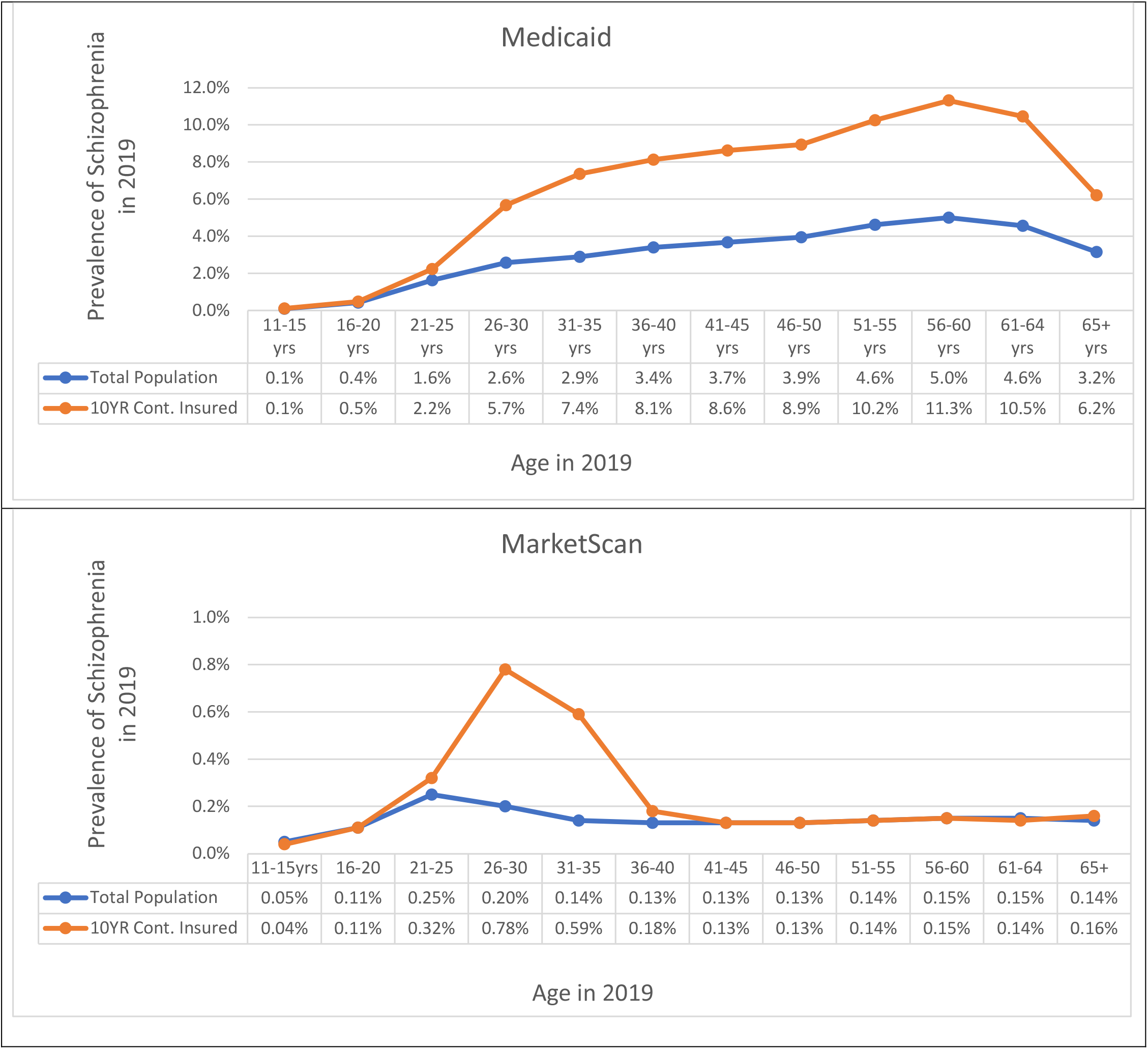
Prevalence of schizophrenia in Medicaid and MarketScan for total population and for those with 10-years continuous insurance by age group over the lifespan (2019)

### Incidence

The incidence of schizophrenia diagnoses in 2019 for the 10-year continuously insured cohort is presented in Table 3. Incidence was higher in Medicaid (0.19%, *n* = 1,489 newly diagnosed in 2019) than in MarketScan (0.07%, *n* = 712), and for males than females (Medicaid: 0.21 vs. 0.18%; MarketScan: 0.08 vs. 0.07%). While incident cases made up only a small proportion of the prevalent cases in the 10-year continuously insured cohort in Medicaid in 2019 (3.3%), incident cases constituted nearly half of prevalent cases in MarketScan (47%).

**Table 3.** Incidence of schizophrenia in Medicaid and MarketScan (2019, 10-years continuously insured)

| **Total, Prevalent and Incident Populations (2019)** | **Incidence in Medicaid** |  |  |  | **Incidence in MarketScan** |  |  |  |
| --- | --- | --- | --- | --- | --- | --- | --- | --- |
|  | **Total** | **Male** | **Female** | **≤35 years** | **Total** | **Male** | **Female** | **≤35 years** |
| Total Insured (*n*) | 785,088 | 305,781 | 479,307 | 302,619 | 951,173 | 361,235 | 589,938 | 220,307 |
| Schizophrenia (*n*) | 45,068 | 24,130 | 20,938 | 4,687 | 1,519 | 664 | 855 | 457 |
| New Onset Schizophrenia (*n*) | 1,489 | 650 | 839 | 493 | 712 | 282 | 430 | 179 |
| **Incidence (%)** | **0.19%** | **0.21%** | **0.18%** | **0.16%** | **0.07%** | **0.08%** | **0.07%** | **0.08%** |
| **Prevalence (%)** | **5.74%** | **7.89%** | **4.37%** | **1.55%** | **0.16%** | **0.18%** | **0.14%** | **0.21%** |
| **Proportion of Incident/**<br>**Prevalent cases (%)** | **3.3%** | **2.7%** | **4.0%** | **10.5%** | **46.9%** | **42.5%** | **50.3%** | **39.2%** |

Incidence by age group is presented in Figure 3. The pattern is similar in both databases, with incidence rising rapidly in late adolescence and peaking in young adulthood (26-30 years of age, Medicaid: 0.40%; MarketScan: 0.16%). New cases of schizophrenia continue to emerge over the lifespan in both populations, but at a lower frequency.

**Figure 3.**
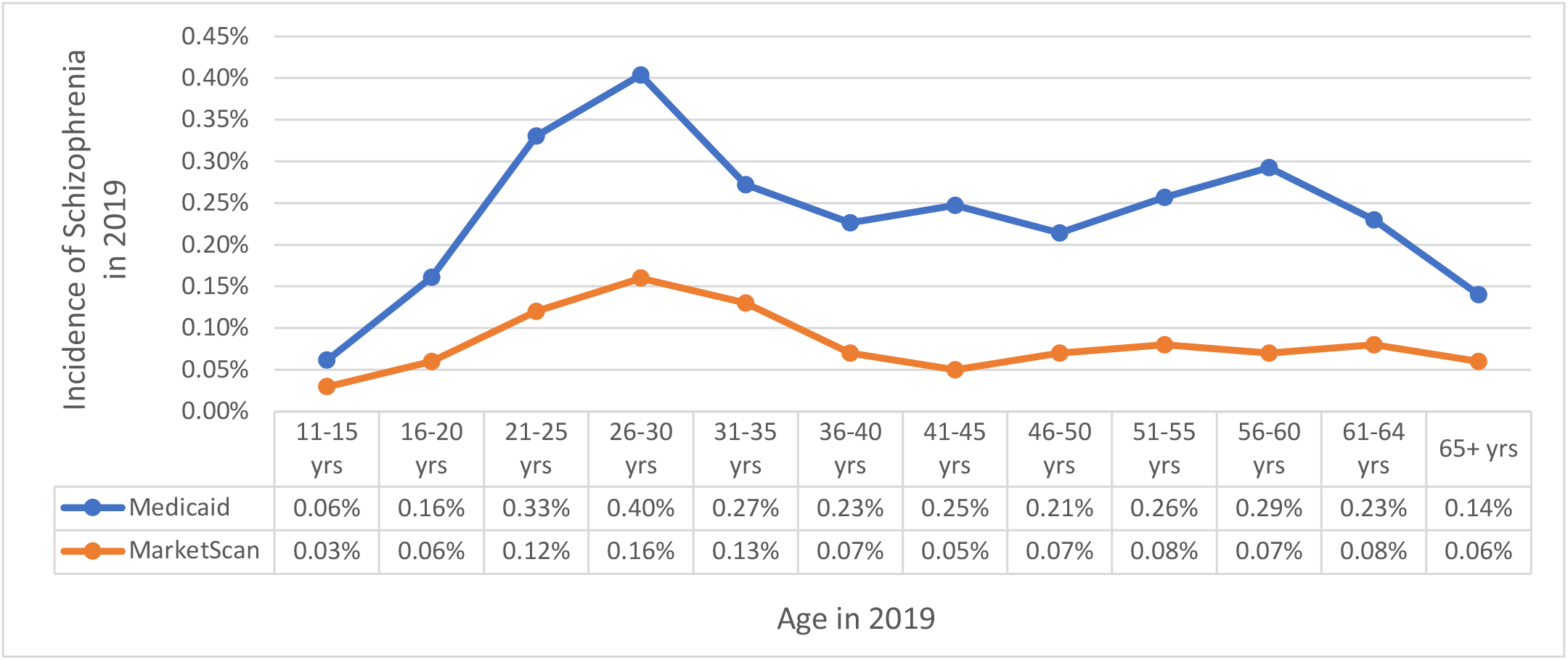
Incidence of schizophrenia in 2019 in Medicaid and MarketScan by age group (10-years continuously insured)

### Accuracy of Incidence Measures

Table 4 summarizes the positive predictive value (PPV) for schizophrenia incidence measures with different periods of observation prior to the first schizophrenia diagnosis in 2019, using a 10-year reference standard. One year of observation prior to the index schizophrenia diagnosis yielded a PPV of 30% for the total Medicaid population, and 51% for those ≤35 years of age. The one-year prior period incidence measure performed better in MarketScan, with a PPV of 81% in the total MarketScan population, and 79% for those ≤35 years of age. Increasing the period of observation prior to the index schizophrenia diagnosis increased the PPVs of the incidence measures in both databases. A PPV of ≥95% was achieved in Medicaid for individuals ≤35 years of age using a minimum of seven years of prior observation, while for MarketScan a minimum period of only two years of prior observation was required. For both Medicaid and MarketScan, PPVs of the incidence measures tested were generally higher for females than for males.

**Table 4.**
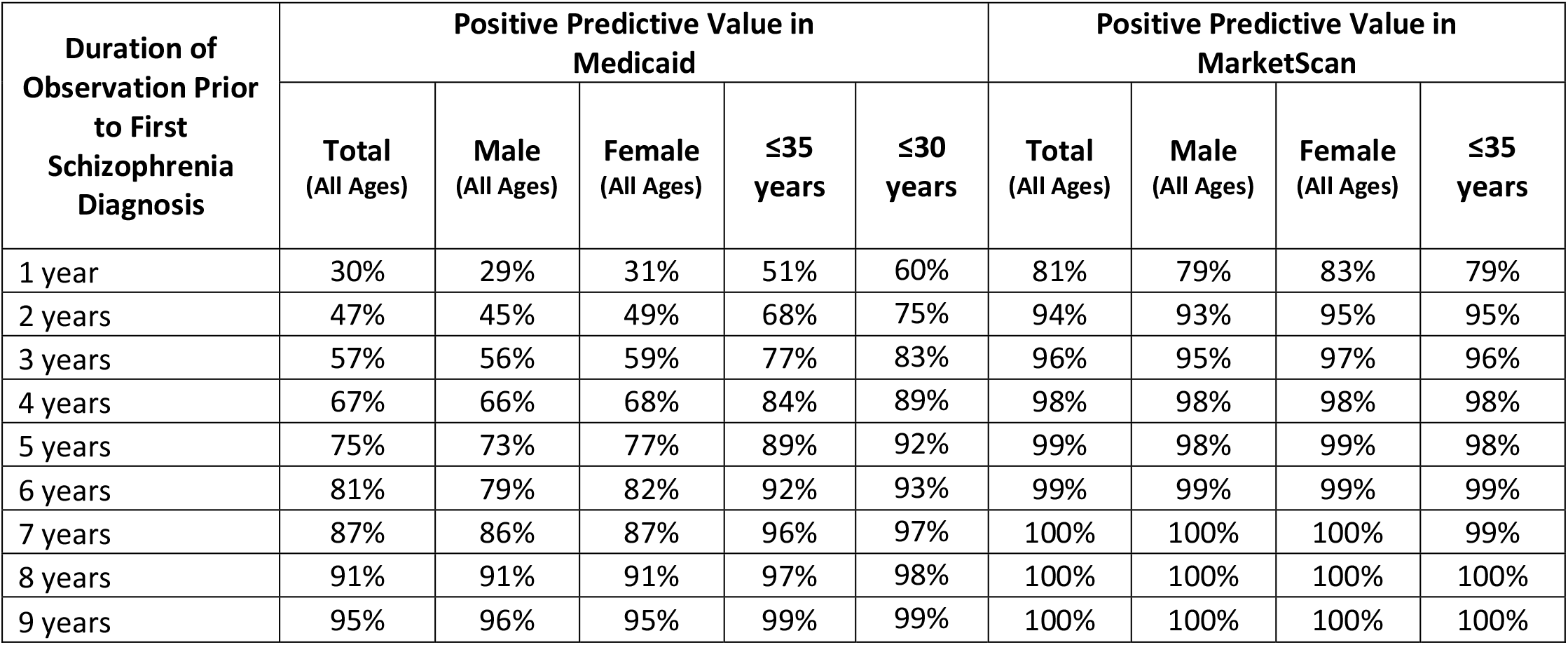
Positive predictive value (PPV) of incidence measures of schizophrenia by duration of observation prior to the index schizophrenia diagnosis in Medicaid and MarketScan populations (2019, 10-years continuously insured)

Figure 4 presents the PPVs of schizophrenia incidence measures by age group and by years of observation prior to the index schizophrenia diagnosis in 2019. In general, PPVs for new-onset schizophrenia measures increase with increasing years of observation prior to the index diagnosis, and are higher for younger age groups. In Medicaid, incidence measures had the highest PPVs for individuals 11-15 years of age, followed by 16-20 years, 21-25 years, 26-30 years, and 31-35 years, with the lowest for individuals 65+ years of age. In MarketScan, the PPVs for incident measures for all age groups were generally higher than Medicaid for all age groups, but the younger age groups performed relatively less well, with the lowest PPV for individuals 26-30 years of age who achieved a 91% PPV by three years, and required seven years of prior observation to achieve a ≥95% PPV.

**Figure 4.**
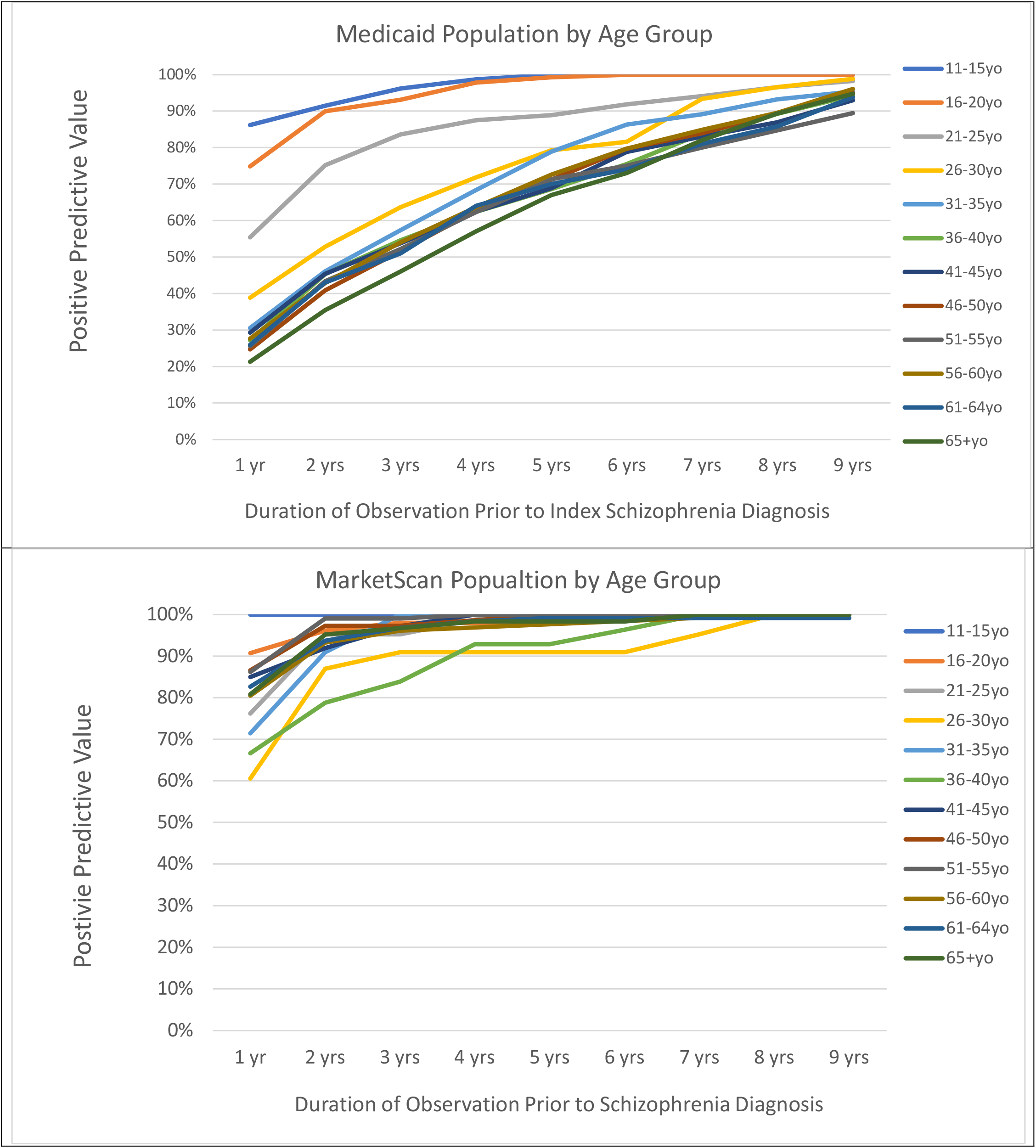
Positive predictive value of schizophrenia incidence measures by duration of observation prior to index schizophrenia diagnosis for different age groups in Medicaid and MarketScan populations (2019, 10-years continuously insured)

The specificity of measures of new-onset schizophrenia was high in both Medicaid and MarketScan, with ≥95% specificity achieved in both databases using two or more years of observation prior to the index schizophrenia diagnosis (Table 5). Specificity was generally higher in Medicaid than in MarketScan (92% vs. 80% for the total population using one year of observation prior to the index schizophrenia diagnosis).

**Table 5.**
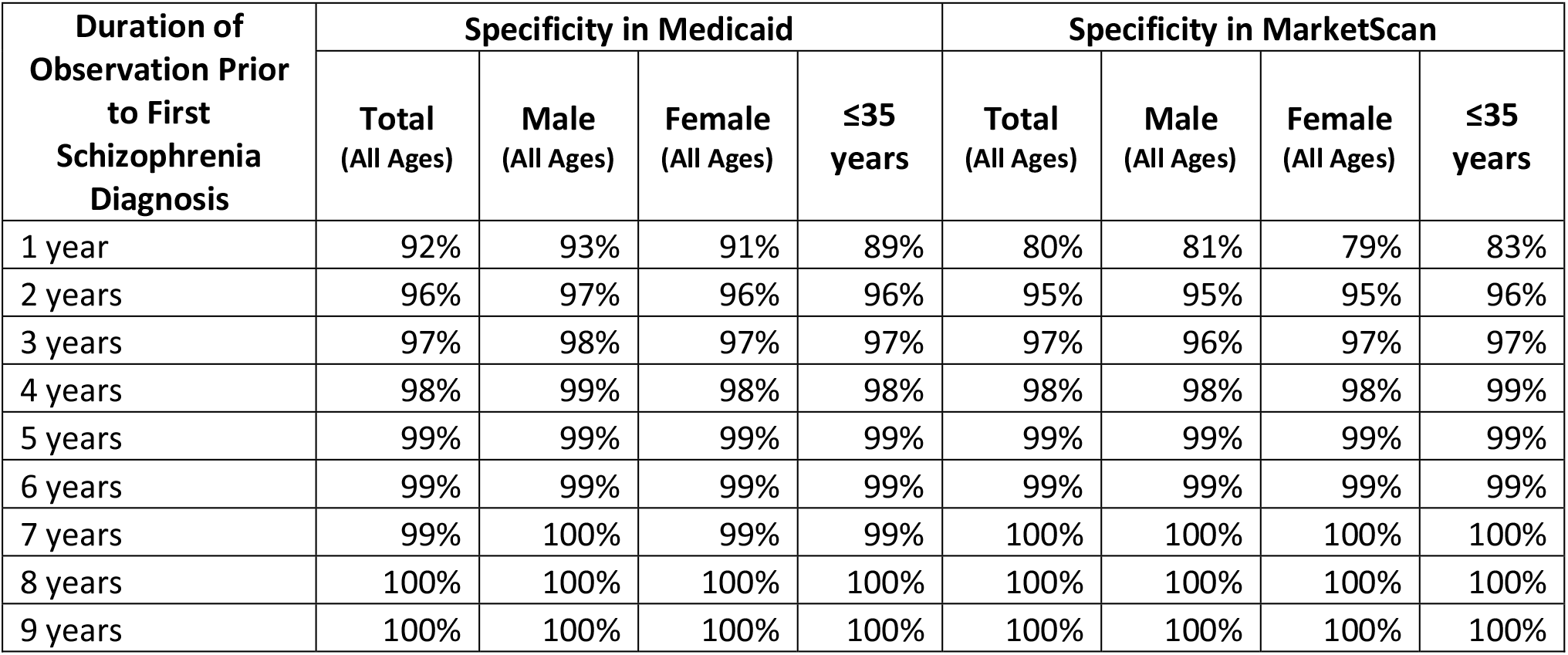
Specificity of incidence measures of schizophrenia by duration of observation prior to index schizophrenia diagnosis in Medicaid and MarketScan populations (2019, 10-year continuously insured)

| Duration of Observation Prior to First Schizophrenia Diagnosis | Specificity in Medicaid |  |  |  | Specificity in MarketScan |  |  |  |
| --- | --- | --- | --- | --- | --- | --- | --- | --- |
|  | Total (All Ages) | Male (All Ages) | Female (All Ages) | ≤35 years | Total (All Ages) | Male (All Ages) | Female (All Ages) | ≤35 years |
| 1 year | 92% | 93% | 91% | 89% | 80% | 81% | 79% | 83% |
| 2 years | 96% | 97% | 96% | 96% | 95% | 95% | 95% | 96% |
| 3 years | 97% | 98% | 97% | 97% | 97% | 96% | 97% | 97% |
| 4 years | 98% | 99% | 98% | 98% | 98% | 98% | 98% | 99% |
| 5 years | 99% | 99% | 99% | 99% | 99% | 99% | 99% | 99% |
| 6 years | 99% | 99% | 99% | 99% | 99% | 99% | 99% | 99% |
| 7 years | 99% | 100% | 99% | 99% | 100% | 100% | 100% | 100% |
| 8 years | 100% | 100% | 100% | 100% | 100% | 100% | 100% | 100% |
| 9 years | 100% | 100% | 100% | 100% | 100% | 100% | 100% | 100% |

## DISCUSSION

This study examined the prevalence and incidence rates of schizophrenia using two administrative databases in the United States and tested the accuracy of incidence measures using varying periods of observation prior to first observed diagnosis by age group. We found a higher prevalence (over 10-fold difference) and incidence (over two-fold difference) of schizophrenia in Medicaid compared to MarketScan. Moreover, the prevalence and incidence were higher in both databases for males compared to females, and among individuals with 10 years or more of continuous insurance compared to those with fewer years of continuous insurance. Accuracy testing identified incidence measures for new diagnoses of schizophrenia with high (≥95%) positive predictive value and specificity in both MarketScan and Medicaid by adjusting the lookback period and age studied. A seven-year period of observation prior to the first documented schizophrenia diagnosis yielded ≥95% positive predictive value for individuals ≤35 years of age, but depending upon the database and the age group under study, periods of prior observation as short as two years were found to be similarly robust.

These findings have implications for surveillance and future research on incidence and outcomes of schizophrenia. Depending on the study question and database used, it may be necessary to restrict the analysis to defined subcohorts, by age and duration of continuous insurance prior to first observed diagnosis of schizophrenia. MarketScan’s commercial insurance database excels at identifying incident cases, even among older adults, as evidenced by the 96% positive predictive value for the total population using a three-year period of observation prior to first schizophrenia diagnosis. In contrast, Medicaid required a seven-year period of prior observation and restriction to younger ages (individuals ≤35 years-old) to achieve a similarly high PPV. However, commercial-based administrative databases in the United States are a less valid measure of prevalence due to the loss of coverage in young adulthood, as evidenced by the peak prevalence we observed in the 26-30 year-old age group and the high proportion of incident to prevalent cases in the total MarketScan population, which would not otherwise be the expected pattern for a chronic disease. The loss of commercial insurance coverage in young adulthood among individuals with schizophrenia mirrors the same trend in the general population (Pottick, Bilder, Vander Stoep, Warner, & Alvarez, 2008), but is greatly exacerbated among individuals with schizophrenia due to chronic unemployment, and the typically young adult-onset of psychosis with progressive cognitive impairment (Gallagher III, 2018). Additional studies are needed to better understand the impact of a schizophrenia diagnosis on retention in commercial insurance.

The polynomial pattern of schizophrenia incidence is demonstrated by elevated rates during young adulthood followed by substantially reduced rates thereafter, with increases during middle age and older adulthood. As proposed by Simeone et al. (2015), examination of prevalence and incidence rates over the lifespan provides a more comprehensive understanding that might otherwise be overlooked in surveillance studies that exclusively focus on young adulthood.

## LIMITATIONS

This study has several limitations. Both MarketScan and Medicaid data are administrative claims databases, for which only those services that are invoiced are recorded. Moreover, administrative health databases rely upon diagnoses assigned in routine clinical practice, not research diagnostic assessments. Notably however, a meta-analysis of the accuracy of mental health diagnoses in administrative data found the highest positive predictive values for the broad category of psychotic illness (PPV ≥80%, with most studies ≥90%) and schizophrenia (PPV ≥75% for the majority of studies), with diagnostic errors occurring primarily at the level of the clinician, rather than errors in administrative data processing (Davis et al., 2016). MarketScan is a national database, predominantly of employed individuals and their families, while the Medicaid database is a large state program, with high rates of unemployment, poverty, and disability. Therefore, it would be expected that some findings may not generalize across databases. However, by incorporating these two distinct populations in our analyses, we reasoned that the likelihood that the resulting measures will generalize to other databases would be enhanced.

## Data Availability

All data produced in the present study are available upon reasonable request to the authors

## CONCLUSION

Incident cases of new diagnoses of schizophrenia can be identified in electronic health record databases with excellent validity for individuals 35 years and younger, with positive predictive values and specificity exceeding 95%. Combining multiple databases that encompass both Medicaid and commercial insurance provides a more comprehensive view of prevalence and incidence of schizophrenia. Further work is needed to leverage these findings to develop robust clinical outcome predictors for diagnosis, prevention, treatment, and prognosis of schizophrenia and other related mental health conditions.

## Notes

Conflicts: All authors report no conflicts of interest specifically related to this work.

### Competing Interest Statement

The authors have declared no competing interest.

### Funding Statement

This study did not receive any funding

### Author Declarations

The Nathan Kline Institute for Psychiatric Research IRB The University of Chicago IRB

### Summary of Updates

This revision has been updated to include Shalmali Joshi and Noemie Elhadad as co-authors.

